# Insulin Resistance Risk in Normal BMI Individuals: Investigating the Role of Genetic Polymorphisms in RNF138, ABCA1, and ESRRG-GPATCH2 Genes - A Case-Control Study in the Indian Population

**DOI:** 10.1101/2024.08.22.24311857

**Authors:** Sabitha Thummala, Sarah Fathima, Nithya Kruthi, Junaid Ahmed Khan Ghori, Katherine Saikia, Vivek Belde, AR Balamurali, Rahul Ranganathan

## Abstract

**Background:** India, characterised as the “diabetes capital” of the world, faces a rapidly increasing diabetes crisis with over 65 million cases diagnosed. Despite the growing prevalence, the genetic underpinnings of insulin resistance (IR) among Indians with normal BMI remain understudied. This research aims to fill the knowledge gap by investigating the association of specific gene variants (*RNF138, ABCA1*, and *ESRRG-GPATCH2*) with IR risk in this demographic.

**Methods:** A total of 191 participants (90 men, 101 women) were recruited for this cross-sectional study. Participants were categorized into cases (HOMA2-IR > 2) and controls (HOMA2-IR < 2) based on Homeostasis Model Assessment Insulin Resistance values. Genotyping for rs4799327 (*RNF138*), rs2275543 (*ABCA1*), and rs1497828 (*ESRRG-GPATCH2*) was performed using the Illumina Infinium Global Screening Array. Statistical analyses, including odds ratios (ORs), 95% confidence intervals (CIs), and inheritance model analysis, were conducted to assess the association between genotypes and IR.

**Results:** Significant associations were found between IR and genetic variants rs4799327 in RNF138 and rs1497828 in *ESRRG-GPATCH2* (dominant inheritance model) and rs2275543 in ABCA1 (additive model). The study highlights a notable susceptibility to IR linked to these genetic markers among normal BMI individuals in the Indian population.

**Conclusions:** This study underscores the importance of genetic factors in the risk of developing insulin resistance among Indians with normal BMI, suggesting a complex interplay of genetics beyond traditional risk factors. These findings necessitate further research into the functional significance of these associations and their potential implications for targeted interventions and preventive strategies in high-risk populations.

**Trial registration:** Not applicable.

## Introduction

The global prevalence of diabetes presents a daunting challenge to public health, marking it as a significant global health crisis. With India accounting for over 77 million diagnosed cases, the nation has emerged as the epicentre of the diabetes epidemic worldwide [1]. The alarming rise in diabetes cases underscores the urgent need for comprehensive research and intervention strategies. India, in particular, is grappling with a staggering rise in diabetes cases. Over the past few decades, the country has witnessed a steady increase in diabetes prevalence, with current estimates indicating over 77 million individuals living with diabetes[3]. Alarmingly, approximately 57% of adults with diabetes in India remain undiagnosed, pointing to significant gaps in healthcare access and screening efforts[1].

Diabetes, as part of the metabolic syndrome, presents a multifaceted challenge in management and treatment. BMI fails to fully capture metabolic nuances; individuals with normal BMI may still have metabolic abnormalities, while those with higher BMIs may exhibit better metabolic health than expected. This limitation underscores the need for comprehensive measures that consider factors like waist circumference, body composition, and metabolic biomarkers to ensure accurate assessment and appropriate management strategies for metabolic disorders like diabetes.[4]

Insulin resistance (IR) stands as a critical precursor to type 2 diabetes, a condition with increasing prevalence worldwide and particularly pronounced in populations like India, where genetic predispositions and lifestyle factors converge to amplify the risk. This concept underscores the intricate interplay between genetic and environmental factors in metabolic dysregulation, highlighting the need for comprehensive research to elucidate the underlying mechanisms. IR, a key factor in metabolic disorders like type 2 diabetes mellitus (T2DM), is closely associated with altered body composition parameters, including increased visceral adiposity or reduced muscle mass, even among individuals with normal Body Mass Index (BMI).[5]

These alterations instigate systemic inflammation and disrupt the balance of adipokines, cytokines, and metabolic signalling molecules, exacerbating insulin resistance. Changes in body composition, accompanied by alterations in metabolic markers such as glycated hemoglobin (HbA1c), fasting plasma glucose (FPG), triglycerides (TG), low-density lipoprotein cholesterol (LDL-C), high-density lipoprotein cholesterol (HDL-C), and total cholesterol (TC), indicate insulin resistance and its associated metabolic disorders, raising the risk of cardiovascular diseases.Through the exploration of genes like *RNF138, ABCA1*, and *ESRRG-GPATCH2*,[6,7,8] crucial in metabolic regulation, this study aims to explore the genetic pathways underlying insulin resistance and its metabolic implications in the Indian population, providing insights into disease susceptibility and potential therapeutic targets.

*RNF138, ABCA1*, and *ESRRG-GPATCH2* have been chosen for investigation in our research due to their recognized involvement in metabolic pathways associated with insulin sensitivity and glucose homeostasis [9].*RNF138*, a type of E3 ubiquitin ligase, is believed to play a role in several cellular functions, including insulin signalling and glucose metabolism, suggesting its potential involvement in the development of IR [10]. Similarly, *ABCA1*, a key regulator of cellular cholesterol efflux, plays a crucial role in lipid metabolism and has been implicated in insulin sensitivity and glucose uptake, making it a promising gene for exploring IR risk [11]. Furthermore, *ESRRG*, a member of the nuclear receptor superfamily, serves as a transcriptional regulator of genes involved in energy metabolism and mitochondrial function, suggesting its potential role in modulating insulin sensitivity and glucose utilisation [12].

By focusing on these genes, the study seeks to elucidate the intricate genetic pathways contributing to IR risk among individuals with normal BMI, thereby enhancing our understanding of the genetic architecture of metabolic dysregulation in the Indian population. Leveraging advanced genetic analyses and statistical methodologies, the study aims to identify specific single-nucleotide polymorphisms (SNPs) within *RNF138, ABCA1*, and *ESRRG-GPATCH2* that are significantly associated with insulin resistance, shedding light on their functional significance and potential implications for diabetes risk.

The current study aims to bridge the gap in our understanding of the genetic factors contributing to IR risk among individuals with normal BMI in the Indian population. By elucidating the genetic underpinnings of IR and its implications for diabetes risk, the study endeavours to inform public health strategies aimed at preventing and managing diabetes in high-risk populations, thereby addressing a critical aspect of the global diabetes epidemic.

## Materials And Methods

### 1. Study Participants

The study involved 191 Indian participants (90 men and 101 women) residing in India, aged between 18 to 65 years, recruited from January 2022 to December 2023. The participants were divided into cases and controls based on their Homeostasis Model Assessment Insulin Resistance (HOMA2-IR) values[13]. Those with HOMA2-IR > 2 were considered cases (57 participants), while those with HOMA2-IR < 2 were regarded as controls (134 participants). HOMA2-IR values were determined using the online tool provided by the Medical Science Division of The University of Oxford[www.OCDEM.ox.ac.uk]. Demographic information, including self-reported height (in centimetres) and weight (in kilograms), was collected through an electronic registration questionnaire. Body mass index (BMI) was computed as weight (in kilograms) divided by height (in metres) squared. Additionally, measurements such as body fat percentage (BFP) and fat mass were obtained. The study excluded individuals with a history of cancer, cardiovascular or renal failure, mental illness, pregnancy, or lactation to ensure participant safety and adhere to ethical guidelines.

### 2. Laboratory Measurements

Following a 12-hour fast, venous blood samples were drawn from participants to assess various metabolic markers. These included glycated haemoglobin (HbA1c), high-density lipoprotein cholesterol (HDL-C), low-density lipoprotein cholesterol (LDL-C), triglycerides (TG), and fasting plasma glucose (FPG).The assessments were conducted using Beckman DxC 700 AU [14] for all markers except HbA1c, which was analyzed with Tosoh G-8[15], and fasting insulin levels measured by Beckman UniCel DxI 800[16], adhering to the protocols provided by the manufacturers,with reference ranges being: FPG (70-100 mg/dl), TC (0-100 mg/dl), TG (0-150 mg/dl), HDL-C (40-60 mg/dl), and LDL-C (0-100 mg/dl). The Homeostasis Model Assessment of Insulin Resistance (HOMA2-IR) formula, which is calculated as fasting insulin (mU/l) x fasting plasma glucose (mmol/l), was used to test insulin resistance[17]. Using a HOMA2-IR cut-off of 2, participants were then classified as either insulin sensitive or resistant. Table 2 lists the clinical traits of the individuals along with how these groupings are associated with them.

**Table 1:** Description of the three single-nucleotide polymorphisms.

| Descriptions for the 3 single-nucleotide polymorphisms |  |  |  |
| --- | --- | --- | --- |
| Gene | Chromosome | RSID | Major/Minor Alleles |
| *RNF138* | 18:29674992 | rs4799327 | G/A |
| *ABCA1* | 9:107651174 | rs2275543 | T/C |
| *ESRRG-GPATCH2* | 1:217527024 | rs1497828 | C/G |
| *RNF138* -ring finger protein 138, *ABCA1*- ATP Binding Cassette Subfamily A Member 1- , *ESRRG-GPATCH2* -estrogen related receptor gamma-G-Patch Domain Containing 2 |  |  |  |

**Table 2:**
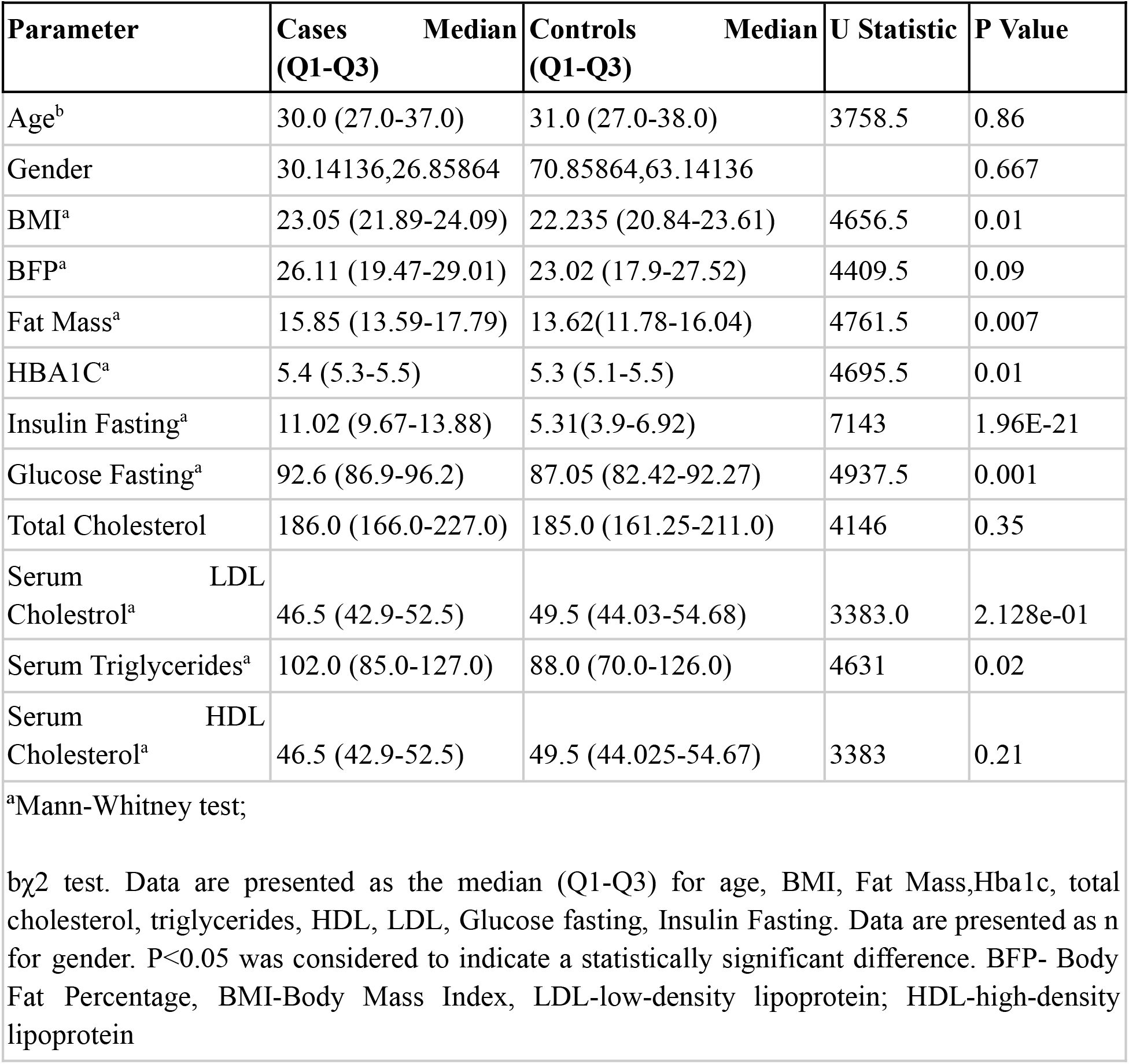
Clinical characteristics and glucose and lipid metabolic parameters of the subjects enrolled in the present study.

### 3. Genotyping and Single-nucleotide polymorphism(SNP) selection

This research aimed to explore the genetic factors contributing to insulin resistance through the examination of participants’ DNA. DNA extraction from blood samples was carried out using the Qiagen blood extraction kit, renowned for its ability to isolate high-quality genomic DNA from blood specimens. The Illumina Infinium Global Screening Array (GSA) V3 platform, renowned for its extensive coverage of genetic variants across the genome, was utilized for the genotyping of single nucleotide polymorphisms (SNPs).The genotyping process utilised the Illumina iScan system, with data interpretation, quality control, and export facilitated by Genome Studio V2 software[18].In this study, we focused on analysing three different genes (*RNF138, ABCA1*, and *ESRRG*) with potential involvement in metabolism and their potential impact on insulin resistance as shown in Table 1. Selection of SNPs was based on an extensive review of scientific literature identifying genetic variations associated with metabolic traits.

The statistical evaluation began by calculating differences in age, glucose levels, and lipid metrics including total cholesterol, LDL cholesterol, HDL cholesterol, triglycerides, and HbA1c between two groups. Due to the non-parametric nature of the data, the analysis employed the Mann-Whitney U test, utilising the Scipy library [19]. Additionally, the distribution of gender across the groups was assessed using the Chi-square test [20], with comprehensive results outlined in Table 2.

### 4. Statistical analysis

The study employed PLINK software for quality control of genotype data, which involved filtering out samples with low genotyping rates, excluding SNPs with considerable missing data, and removing rare variants due to their limited statistical power.Heterozygosity checks were performed to verify the accuracy of genotype distribution. Following quality control, SNPstats software was utilised to investigate the association between specific SNPs and insulin resistance[21]. The analysis produced odds ratios (ORs) along with their corresponding 95% confidence intervals (CIs). Table 3 presents the findings, highlighting significant SNP markers across various genetic inheritance models.

**Table 3:** The risk alleles for the polymorphisms along with the best fit model and statistical significance.

|  |  |  |  |  |
| --- | --- | --- | --- | --- |
| Gene | RSID | Risk Allele | Best Fit Model | Statistical Significance |
| RNF138 | rs4799327 | A | Dominant | (OR)=2.74; 95%(CI): 1.28-5.88; P = 0.006 |
| ABCA1 | rs2275543 | C | Log additive | (OR)=3.50; 95%(CI): 1.17-10.42; P = 0.011 |
| ESRRG-GPATCH2 | rs1497828 | C | Dominant | (OR)=2.90; 95%(CI):1.51-5.57; P =0.0011 |

**Table 4:** Comparison of genotypic and allelic distribution of three SNPs (rs4799327, rs2275543 and rs1497828) between the two groups HOMA2 IR >2 and HOMA2 IR <2.

| **Table 4: Comparison of Allelic and Genotypic Distribution of three SNPs (rs4799327, rs2275543 and rs1497828) between Cases and Controls** |  |  |  |  |  |  |  |  |  |  |
| --- | --- | --- | --- | --- | --- | --- | --- | --- | --- | --- |
| **SNP** | **Group** | **A1** | **A2** | **$\chi^2$** | **$\chi^2$**<br>**P-Value** | **A1A1** | **A1A2** | **A2A2** | **P-Value** | **HWE**<br>**P-Value** |
| **rs4799327** | cases | G<br>102<br>(0.89) | A<br>12<br>(0.11) | 5.182 | 0.0228<br>2 | A/A<br>1<br>(0.02) | G/A<br>10<br>(0.18) | G/G<br>46<br>(0.81) | 0.0063 | 0.48 |
|  | controls | G<br>214<br>(0.8) | A<br>54<br>(0.2) |  |  | A/A<br>4<br>(0.03) | G/A<br>46<br>(0.34) | G/G<br>84<br>(0.63) |  | 0.59 |
| **rs2275543** | cases | T<br>110<br>(0.96) | C<br>4<br>(0.04) | 5.017 | 0.0250<br>9 | C/C<br>0<br>(0) | T/C<br>4<br>(0.07) | T/T<br>53<br>(0.93) | 0.011 | 1 |
|  | controls | T<br>240<br>(0.9) | C<br>28<br>(0.1) |  |  | C/C<br>1<br>(0.01) | T/C<br>26<br>(0.19) | T/T<br>107<br>(0.8) |  | 1 |
| **rs1497828** | cases | C<br>27<br>(0.24) | G<br>87<br>(0.76) | 9.264 | 0.0023<br>38 | C/C<br>4<br>(0.07) | G/C<br>19<br>(0.33) | G/G<br>34<br>(0.6) | 0.0013 | 0.053 |
|  | controls | C<br>107<br>(0.4) | G<br>161<br>(0.6) |  |  | C/C<br>18<br>(0.13) | G/C<br>71<br>(0.53) | G/G<br>45<br>(0.34) |  | 0.73 |
| **Note: In the chi-square association analysis of cases and controls, the four SNPs (rs920590, rs7274134, rs6762208 and rs2078267) exhibited statistically significant associations with the trait after Bonferroni correction, with their respective p-values falling below the adjusted threshold of 0.016.** |  |  |  |  |  |  |  |  |  |  |

**Table 4.**
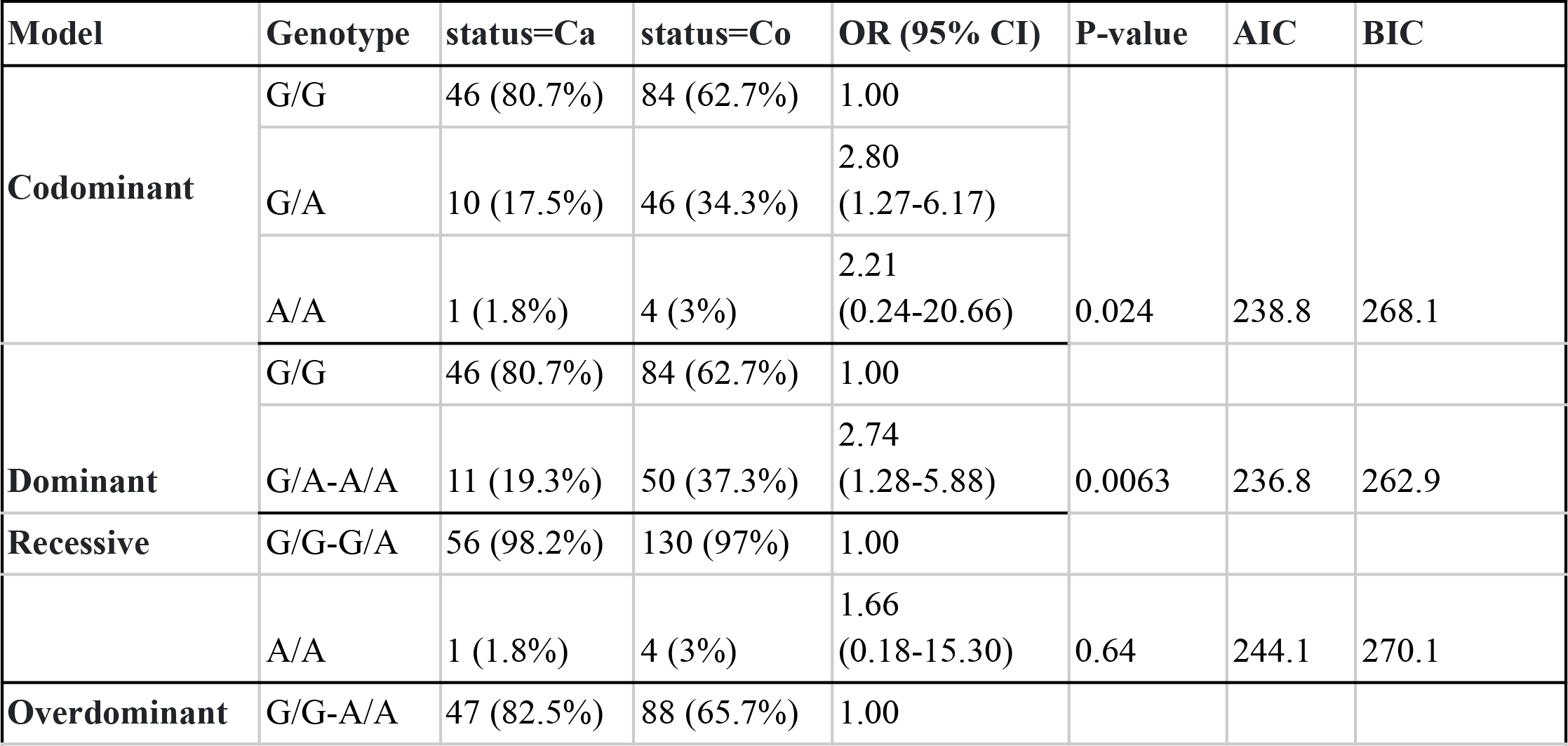
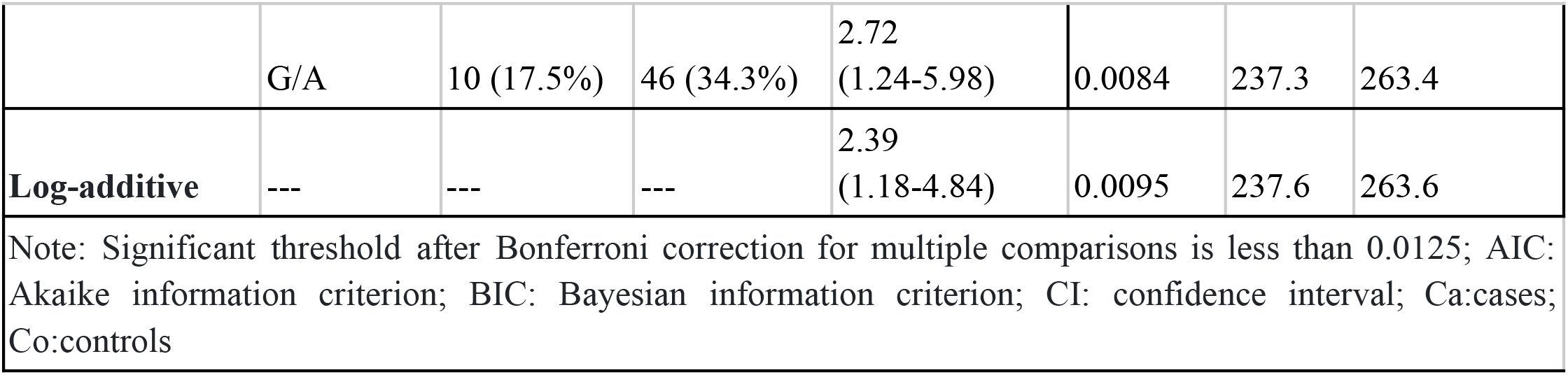
Inheritance models analysis of the SNP rs4799327 (RNF198) between the Cases and Controls rs4799327 association with response status (n=191, adjusted by gender+age.cat.

In the control group, adherence to Hardy-Weinberg equilibrium was maintained at a significance level of 0.05[21]. Calculating odds ratios (OR) and the associated 95% confidence intervals (95% CI) was the method used to estimate risk. The linkage disequilibrium (LD) between the SNPs was also investigated in the study, using the LD coefficient D that was determined using the Snpstats software. Additionally, power analyses were performed based on power and sample size calculations[22]. Each SNP underwent Bonferroni adjustment to mitigate the risk of type I errors resulting from multiple tests.

## Results

### Subject characteristics

In this study, a total of 191 participants were included, and all underwent genotyping. The clinical characteristics, as well as glucose and lipid metabolic parameters of the subjects, are detailed in Table 2. The analysis conducted on the data did not reveal any significant statistical differences in terms of age or gender between the case and control groups, nor among males and females within those groups (p > 0.05). However, notable variations were observed in BMI (p = 0.01), Fat Mass (p = 0.007), HbA1c (p = 0.01), Fasting Glucose (p = 0.001), and Serum Triglycerides (p = 0.02) when comparing cases to controls. It’s worth mentioning that exceptions were noted for Body Fat Percentage (BFP), Total Cholesterol, and Serum HDL Cholesterol.

### Allelic and Genotypic Analysis

The Allelic and genotypic distribution of the three SNPs are given in the below Table 3. All the controls and cases for the SNPs rs4799327, rs2275543 and rs1497828 are in compliance with the Hardy Weinberg equation (p>0.05).This was evaluated through a Fisher’s exact test.

Chi-square tests were performed to evaluate the association between genotypes and allele frequencies of each SNP with phenotype (IR). All the SNPs show significant differences between the cases and control after the Bonferroni Correction with respective p values as 0.0063, 0.011 and 0.0013 for the Genotypic frequencies (p<0.016). Thus, the association between these three polymorphisms and Insulin Resistance is quite significant. The Allelic frequency difference between the cases and controls for all the SNPs in consideration also stands significant (p< 0.05).

Chi-squared analyses were performed to investigate the relationship between the genotypic and allelic variations of four SNPs (rs4799327, rs2275543 and rs1497828) within two groups distinguished by their HOMA2 IR levels (>2 and <2).The obtained p-values indicate significant associations between SNP genotypes and insulin resistance status.

The risk alleles for the polymorphisms are illustrated in the given table:

### Model Of Inheritance Analysis

In this study, genetic inheritance models (codominant, dominant, recessive, overdominant, and log-additive models) for SNPs rs4799327, rs2275543, and rs1497828 were investigated. Tables 5 - 7 provide detailed data on these models, including the evaluation of the lowest Akaike Information Criterion (AIC) and Bayesian Information Criterion (BIC) values to determine the best-fit model for each SNP[23].

**Table 5.** Inheritance models analysis of the SNP rs2275543 (ABCA1) between the Cases and Controls rs2275543 association with response status (n=191, adjusted by gender+age.cat)

| **Table 5 Inheritance models analysis of the SNP rs2275543 (ABCA1) between the Cases and Controls** |  |  |  |  |  |  |  |
| --- | --- | --- | --- | --- | --- | --- | --- |
| **rs2275543 association with response status (n=191, adjusted by gender+age.cat)** |  |  |  |  |  |  |  |
| Model | Genotype | status=Ca | status=Co | OR (95% CI) | P-value | AIC | BIC |
|  | T/T | 53 (93%) | 107 (79.8%) | 1.00 |  |  |  |
| **Codominant** | T/C | 4 (7%) | 26 (19.4%) | 3.39<br>(1.12-10.30) | 0.036 | 239.7 | 268.9 |
|  | C/C | 0 (0%) | 1 (0.8%) | NA<br>(0.00-NA) |  |  |  |
| **Dominant** | T/T | 53 (93%) | 107 (79.8%) | 1.00 |  |  |  |
|  | T/C-C/C | 4 (7%) | 27 (20.1%) | 3.54<br>(1.17-10.75) | 0.012 | 238 | 264 |
| **Recessive** | T/T-T/C | 57 (100%) | 133 (99.2%) | 1.00 |  |  |  |
|  | C/C | 0 (0%) | 1 (0.8%) | NA<br>(0.00-NA) | 0.36 | 243.4 | 269.5 |
| **Overdominant** | T/T-C/C | 53 (93%) | 108 (80.6%) | 1.00 |  |  |  |
|  | T/C | 4 (7%) | 26 (19.4%) | 3.35<br>(1.10-10.16) | 0.018 | 238.6 | 264.7 |
| **Log-additive** | — | — | — | 3.50<br>(1.17-10.42) | 0.011 | 237.8 | 263.8 |

Note: Significant threshold after Bonferroni correction for multiple comparisons is less than 0.0125; AIC: Akaike information criterion; BIC: Bayesian information criterion; CI: confidence interval; Ca:cases; Co:Controls

**Table 5.** Inheritance models analysis of the SNP rs1497828 (ESRRG-GPATCH2) between the Cases and Controls rs1497828 association with response status (n=191, adjusted by gender+age.cat)

| **Table 5 Inheritance models analysis of the SNP rs1497828 (ESRRG- GPATCH2) between the Cases and Controls** |  |  |  |  |  |  |  |  |
| --- | --- | --- | --- | --- | --- | --- | --- | --- |
| **rs1497828 association with response status (n=191, adjusted by gender+age.cat)** |  |  |  |  |  |  |  |  |
| Model | Genotype | status=Ca | status=Co | OR (95% |  | P-value | AIC | BIC |

Genetic inheritance analysis involves assessing how genetic variants contribute to the inheritance patterns of certain traits or diseases within populations. Different genetic inheritance models are evaluated to understand the relationship between genotypes and phenotypes. These models include codominant, dominant, recessive, overdominant, and log-additive models. The choice of the best-fit model is determined based on statistical criteria such as AIC and BIC values, with lower values indicating a better fit.

For SNP rs4799327 located in the RNF138 gene and rs1497828 in the intergenic region of *ESRRG-GPATCH2*, the top model identified was Dominant, with respective p values of 0.0063 and 0.0011, accompanied by the lowest AIC and BIC values.

Specifically, for SNP rs4799327, the risk genotype was identified as G/A-A/A compared to G/G, exhibiting a significant association (P=0.0063; OR=2.74; 95% CI: 1.28-5.88), as outlined in Table 5.

Regarding SNP rs2275543, the log-additive model emerged as the best fit, characterised by the lowest AIC and BIC values. The risk allele for this model was determined to be C (OR=3.50; 95% CI: 1.17-10.42; P=0.011), as depicted in Table 6.

Finally, for SNP rs1497828, the risk genotype was identified as G/C-C/C in contrast to G/G, displaying a significant association (P=0.0011; OR=2.90; 95% CI: 1.51-5.57), as shown in Table 7.These findings provide valuable insights into the genetic basis of insulin resistance, shedding light on the specific polymorphisms associated with increased susceptibility to this metabolic condition.

## Discussion

Understanding the genetic underpinnings of insulin resistance (IR) and its association with metabolic disorders is crucial for advancing our knowledge of disease mechanisms and developing targeted interventions. In this discussion, we delve into the genetic factors identified in our study specifically, the roles of RNF138, ABCA1, and ESRRG-GPATCH2 polymorphisms in influencing insulin sensitivity and glucose homeostasis. By elucidating the functional significance of these genetic variants and their implications for metabolic health, we aim to contribute to the broader understanding of IR pathogenesis and its potential therapeutic interventions.

RNF138 belongs to the RING finger (RNF) family. And amongst them it belongs to the UIM subfamily in particular [24]. The RING finger proteins contain the RING domain that has E3 ubiquitin ligase which is important for carrying out a highly specific post translational modification known as ubiquitination [25]. E3 ubiquitin ligase is known to contribute to insulin resistance due its proteolytic and degradation effects on the insulin receptor and insulin receptor substrates Bai et al [26]. X-D Yang *et al*., proposes two methods by which E3 ubiquitin ligase affects insulin resistance and Diabetes-one where Insulin receptor substrate, insulin receptor, and other important insulin signalling molecules are directly degraded by E3 ubiquitin ligases via the ubiquitin-proteasome system and second way is where E3 indirectly functions and regulates insulin signalling through the regulation of pro-inflammatory mediators such as TNF-α [6].

In a murine knock out model study by Bhagwandin C et al., it was concluded that MARCH1 which is a RING finger member exhibits a negative regulatory effect on the insulin receptors through ubiquitination, which eventually effects the insulin sensitivity[27]. Hu X et al. found that RNF186 increased hepatic triglycerides buildup, dysregulated insulin sensitivity and could increase hepatic inflammation when on a high fat diet while Lee J H et al., found RNF20 regulates liver triglyceride synthesis[28].RNF213 is linked to adipogenesis and insulin control, according to Sarkar P et al.’s research [29], which connects TNF-mediated pathways to obesity. According to the dominant model of inheritance, our research shows a strong correlation between insulin resistance and rs4799327 in the RNF138 gene (OR)=2.74; 95%(CI): 1.28-5.88; P = 0.006.

The ATP-binding cassette transporter ABCA1 is essential for the transfer of cellular cholesterol and phospholipids to lipid-poor apolipoproteins, which forms precursor high-density lipoprotein (HDL) particles and aids in lipid metabolism.[30] The expression of ABCA1 in adipose tissue and its relationship to insulin resistance (IR) in obesity were studied by Vincent et al. [31]. Their findings showed that obese people had substantially reduced levels of ABCA1 expression in their visceral adipose tissue. Reduced ABCA1 expression was seen in insulin-resistant individuals, and this was independently linked to increased insulin sensitivity.

The review by [32] highlights the multifaceted role of ATP-binding cassette transporter A1 (ABCA1) in metabolic syndrome (MetS), emphasising its involvement in HDL and VLDL production, insulin-glucose homeostasis, inflammation suppression, and obesity regulation. It elucidates how ABCA1 mediates key steps in HDL biogenesis, impacts VLDL production in the liver, and influences insulin secretion and sensitivity in pancreatic β-cells, adipocytes, and skeletal muscle cells. Furthermore, it underscores the association between abnormal ABCA1-regulated phenotypes and the development of MetS, offering insights into potential therapeutic strategies targeting ABCA1 to mitigate MetS-related complications. Our study complements these findings, revealing a significant association between the SNP rs2275543 in the ABCA1 gene and insulin resistance. The identification of the risk allele C (OR=3.50; 95% CI: 1.17-10.42; P = 0.011) further accentuates ABCA1’s role in metabolic disorders, aligning with the review’s emphasis on ABCA1’s involvement in insulin-glucose homeostasis, underscoring its potential as a therapeutic target for addressing MetS-related complications.

De Haan et al.[33] examined how the absence of ABCA1 specifically in adipocytes affects lipid metabolism and glucose regulation using ABCA1 mice. Their research showed that without ABCA1, there was a buildup of cholesterol and triglycerides in adipose tissue, leading to larger fat deposits and increased body weight when fed a diet high in fat and cholesterol. Additionally, ABCA1-deficient mice displayed impaired glucose tolerance, reduced sensitivity to insulin, and lower insulin production. These results emphasise the importance of adipocyte ABCA1 in controlling lipid levels and glucose metabolism, suggesting it could be a promising target for treating metabolic disorders..Our study reveals a significant association between insulin resistance and ABCA1 gene’s SNP rs2275543. The risk allele C is linked to increased insulin resistance (OR=3.50; 95% CI: 1.17-10.42; P = 0.011), shedding light on its potential role in metabolic disorders like type 2 diabetes mellitus.

The estrogen-related receptor γ (ESRRG) is part of the orphan nuclear hormone receptor family, which includes steroid hormone receptors. It acts as a continuous transcription[39] activator. This group of receptors is involved in various regulatory functions, managing both homeostatic and metabolic activities.[34,35]ESRRG is implicated in the pathophysiology related to both the pancreas and the liver. In the pancreas, ESRRG is involved in the regulation of pancreatic β-cell function and insulin secretion, affecting glucose homeostasis. In the liver, ESRRG influences hepatic glucose production and lipid metabolism, contributing to insulin sensitivity and glucose utilisation. Dysregulation of ESRRG signalling pathways can disrupt insulin action and exacerbate insulin resistance, thereby playing a role in the development and progression of metabolic disorders like type 2 diabetes mellitus. GPATCH2, which stands for G-Patch Domain Containing 2, is a gene that encodes a protein known to be involved in RNA processing, an essential step in the production of proteins from genes. While GPATCH2 is not directly implicated in the typical pathways associated with insulin signalling or glucose metabolism, its role in cellular processes might indirectly influence factors related to insulin resistance or the development of diabetes.[36]

The association between the ESRRG rs1890552 A>G polymorphism and urine 8-epi-PGF2α levels was investigated in a study by Kim et al.[37] involving 1933 Korean individuals to see whether it was related to impaired fasting glucose (IFG) or the onset of type 2 diabetes (T2D). The results showed that, relative to the control group, those with IFG or T2D had higher plasma malondialdehyde (MDA), urinary 8-epi-PGF2α, and brachial-ankle pulse wave velocity (baPWV). Furthermore, there was a significant increase in the frequency of the ESRRG rs1890552 GG genotype in individuals with T2D or IFG, suggesting that this genotype may contribute to an increased vulnerability to these metabolic disorders.Our study identified a significant association between SNP rs1497828 and T2D risk, with the G/C-C/C genotype showing higher risk compared to G/G genotype carriers (P=0.0011; OR=2.90; 95% CI: 1.51-5.57). This emphasizes the importance of genetic variations in predisposing individuals to T2D and the utility of genetic markers in identifying those at risk for metabolic disorders.

The rationale behind selecting RNF138, ABCA1, and ESRRG-GPATCH2 lies in their well-established roles in metabolic pathways implicated in insulin sensitivity and glucose homeostasis, making them compelling genes for genetic studies focused on IR risk in the Indian population. Furthermore, understanding the genetic determinants of IR holds significant implications for public health intervention and personalised medicine approaches, paving the way for targeted therapeutic interventions aimed at mitigating the burden of diabetes in high-risk populations.

## Conclusion

In conclusion, our study indicates that the RNF138, ABCA1, and ESRRG-GPATCH2 play significant roles in the genetic predisposition to insulin resistance within the Indian population. Through meticulous examination of relevant SNPs and comprehensive analysis of metabolic markers, we identified notable associations between genetic variations and insulin resistance status. Specifically, our findings reveal compelling links between certain genotypes and increased risk of insulin resistance, shedding light on the intricate genetic architecture underlying metabolic dysfunction. Moreover, our study underscores the importance of further research to validate and extend these findings in larger, prospective longitudinal cohorts. By deepening our understanding of the genetic determinants of insulin resistance, our study lays a crucial foundation for future investigations aimed at developing targeted interventions and personalised therapeutic approaches to mitigate the burden of diabetes and related metabolic disorders in high-risk populations.

## Data Availability

The data will be available upon request to the corresponding author

https://www.nugenomics.in/

## Declarations

### Ethics approval and consent to participate

This study was approved by the Answergenomics Ethical Review Committee Board. We have obtained written consent from all the subjects.

### Consent for publication

Not applicable

### Availability of data and materials

The datasets generated and/or analyzed during the current study are not publicly available due to the sensitive nature of genetic data and privacy concerns for participants. However, data are available from the corresponding author upon reasonable request and with the assurance that the privacy of participants will be maintained.

### Authors’ contributions

ST spearheaded the research efforts, providing invaluable expertise in genetic analysis and study design. SF contributed to Data Analysis, interpretation and drafting of manuscript. NK statistically analysed the genetic data and majorly drafted and reviewed the manuscript. JG analysed large scale genetic dataset, providing genetic associations and complex genetic patterns. KS contributed to the editing and proofreading of the script.VB contributed his clinical insights by offering valuable perspectives. BR and RR conceptualised the study idea and provided the platform to carry out the research.

## Acknowledgements

Not Applicable

## Authors’ information

Answergenomics Pvt Ltd, Delhi, India

## Abbreviations

IR: Insulin Resistance
BMI: Body Mass Index
HOMA2-IR: Homeostasis Model Assessment Insulin Resistance
SNP: Single-+
RNF138: Ring Finger Protein 138
ABCA1: ATP Binding Cassette Subfamily A Member 1
ESRRG-GPATCH2: Estrogen Related Receptor Gamma-G-Patch Domain Containing
OR: Odds Ratio
CI: Confidence Interval
LDL-C: Low-Density Lipoprotein Cholesterol
HDL-C: High-Density Lipoprotein Cholesterol
TC: Total Cholesterol
TG: Triglycerides
FPG: Fasting Plasma Glucose
HbA1c: Glycated Haemoglobin
BFP: Body Fat Percentage
T2DM: Type 2 Diabetes Mellitus
AIC: Akaike Information Criterion
BIC: Bayesian Information Criterion

